# Neighborhood deadwood and yard rewilding modulate commensal microbiota and inflammatory signals among urbanites

**DOI:** 10.1101/2024.09.26.24314419

**Authors:** Marja I. Roslund, Laura Uimonen, Laura Kummola, Damiano Cerrone, Ann Ojala, Anna Luukkonen, Ella Holopainen, Aku Korhonen, Reijo Penttilä, Martti Venäläinen, Hanna Haveri, Juho Rajaniemi, Olli H. Laitinen, Aki Sinkkonen, the BIWE research group

## Abstract

**Background:** Global ecosystem deprivation is linked to reduced microbial diversity and diminished immunological resilience. Urban rewilding and decomposing plant matter have been suggested to reverse this deprivation and support human health.

**Methods:** We rewilded 21 urban private yards with vegetation and deadwood. Control yards (15) were analyzed for comparison. We collected microbial samples and used vegetation and deadwood inventories, landcover data and questionnaires to determine the effects of rewilding and living environment on skin and salivary microbiota, gene pathways and cytokine levels (IL-6, IL-10). Samples were collected before the rewilding in summer and three months later in autumn.

**Findings:** Rewilding preserved skin microbial richness in comparison to control group, including previously health-associated Alpha- and Gammaproteobacteria, despite the normal seasonal decline and less outdoor time in autumn. Deadwood abundance within 200-m radii associated directly to beta diversity of skin microbiota and Gammaproteobacterial taxonomies.

In saliva, deadwood was directly associated with the diversity of functional gene pathways, which in turn was negatively associated with pleiotropic IL-6 levels. Rewilding was associated with a decrease in L-histidine degradation and an increase in Mycobacteriaceae.

**Interpretation:** Since both yard rewilding and neighborhood deadwood preserved rich commensal microbiota and reduced pro-inflammatory signals, decomposing plant matter, not just plant richness, seems to be crucial for ecosystem services that contribute to health. Since deadwood abundance was associated to reduced pro-inflammatory signals, it may be a suitable indicator of environment supporting immunological resilience.

Our findings provide an incentive for future strategic investments for planetary health.

**Funding:** Strategic Research council Finland.

## 1. Introduction

According to current understanding, biodiversity loss and the lack of microbial exposure in urban ecosystems is a core reason for impaired immune system regulation and the epidemic of immune-mediated diseases among urban populations.^1,2^ Hence, there is an unmet need to rewild the urban ecosystems.^3–5^ Urban rewilding has many benefits including increased resilience to environmental change and opportunities for urban dwellers to reconnect with nature.^6^ Urban, private yards contribute remarkably to local ecology, biodiversity and wellbeing of the residents.^7,8^ Microbially diverse private yards are generally assumed to provide ecosystem services that alleviate microbial deprivation and proinflammatory responses among residents,^1,5,9^ however, this has remained untested.

Environmental microbial and plant community diversity influence skin and salivary microbiota.^2,10–18^ Recent studies demonstrated how rewilding of urban daycare yards diversify children’s skin microbiota and enhance immune regulation.^12,13^ The core reason for the enhanced immune regulation seems to be the exposure to diverse microbiome *per se*; when study subjects were exposed to microbially rich sandbox sand^19^ or growing media with high microbial diversity,^15^ immune regulation was enhanced only in the intervention arms, not in placebo groups.

Studies exploring human immune system have frequently used plant diversity indices or plant species richness as an indicator of exposure to microbiological diversity.^2,20–23^ Since a recent review summarized the results of such studies as conflicting,^21^ differences in vegetation diversity hardly provide a complete explanation for the difference in services provided by urban and rural ecosystems.

Since the accumulation of decaying plant debris is recognized as an important driver of biodiversity, the importance of decomposing plant matter could provide an alternative explanation why urban ecosystems seldom support immune modulation.^24^ Indeed, decaying wood is recognized to provide habitats for numerous fungi, saproxylic invertebrates and prokaryotes, particularly bacteria,^25,26^ and people living in environments hosting the rich microbiota have a low probability of certain immune-mediated diseases.^2,27^ Thus, the evaluation of ecosystem services provided by private yards should include indicators of decaying plant matter, e. g., deadwood and wood-inhabiting polypore fungi.^28,29^ In parallel with proper measures of biodiversity, measures of immune function must be feasible and reflect environmental exposure to potential modifiers. Salivary cytokine levels reflect the environmental exposure and are a noninvasive alternative to blood cytokines as biomarkers for inflammatory responses and, neuroimmune function.^30–32^

The effect of rewilding of private yards, including alterations in garden management practices, on health-associated commensal microbiota and salivary cytokine levels has never been tested in an intervention trial. We rewilded urban private yards with diverse vegetation and decaying deadwood and plant residues. We used satellite data and vegetation, deadwood, and polypore inventories to determine the living environment of the participants and biodiversity around home yards. Questionnaires were used to evaluate the yard management practices and living habits of yard owners. Based on previous comparative studies^2,14,33,34^ and intervention trials,^12,13,15,19^ we hypothesized that yard rewilding diversifies health-associated skin microbiota, particularly alpha- and gammaproteobacterial communities, and that the rewilding is associated with salivary cytokine levels, gene pathways and microbiota. We also hypothesized that plant species richness and the abundance of deadwood are indicators of microbial exposure and thus associated with commensal microbiota.

## 2. Materials and Methods

### 2.1. Study group

Thirty-six volunteers aged between 33–70 participated in the study (Table 1). The participants were recruited to two groups. Intervention group, hereafter called as the rewilding group, consisted of volunteers who gave a permission to diversify yard herbaceous and woody species community and were interested in following the yard management instructions described below in section 2.3. Control group continued gardening as they had done before. Study subjects were recruited from April 2022 to June 2022 in three metropolitan regions in Finland; Helsinki metropolitan region (660,000 inhabitants) lies on the south coast of Finland, Lahti (120,000 inhabitants) 100 km north, and Turku (480,000 inhabitants) 150 km west of Helsinki. All these urban housing areas face challenges in preserving green space amid built environment.

**Table 1.** Characteristics of the study participants. Age and land cover categories around home yards with radius 500m are presented as mean ± standard deviation. The land cover categories surrounding the private yards were analyzed with the preclassified Coordination of Information on the Environment (CORINE).

|  | **Intervention** | **Control** | **Total** |
| --- | --- | --- | --- |
| **Gender, male** | 5 | 4 | 9 |
| **Gender, female** | 16 | 11 | 27 |
| **Gender, non-binary** | 0 | 0 | 0 |
| **Age** | 46 ± 11 | 52 ± 13 | 48 ± 12 |
| **Pet ownership** | 12 | 3 | 15 |
| **Land cover categories 500-m radius:** |  |  |  |
| **Artificial surfaces%:** | 94 ± 16 | 95 ± 7 | 95 ± 13 |
| - **Discountinous urban fabric%** | 59 ± 21 | 70 ± 16 | 64 ± 20 |
| - **Industrial or commercial units** | 26 ± 20 | 20 ± 21 | 24 ± 20 |
| - **Green urban areas%** | 9 ± 16 | 3 ± 10 | 6 ± 14 |
| **Forests%** | 2 ± 8 | 1 ± 5 | 2 ± 6 |
| **Agricultural areas%** | 3 ± 13 | 1 ± 4 | 3 ± 10 |

The medical exclusion criteria included immune deficiencies, a disease affecting immune response, cancer diagnosis within the last year or on-going cancer treatment. Other exclusion criteria included incompetency and living outside city area. Inclusion criteria was that the form of housing in a detached or row house. All participants provided a written, informed consent in accordance with the Declaration of Helsinki.

The study strictly followed the recommendations of the “Finnish Advisory Board on Research Integrity”, and it received an ethical approval from the HUS Committee on Medical Research Ethics (Helsinki University Hospital District). The trial has been registered in ClinicalTrials.gov (ID NCT06353035).

The participants filled out electronic questionnaires covering detailed background information, diet, nature visits, medication use, yard management practices, and details about yard activities (Appendix p 3).

### 2.2. Outcome measures and sample size

A primary measure for sample size was the difference on skin gammaproteobacterial diversity between the rewilding and control group. Secondary outcome measures included differences in bacterial richness and diversity (beta and alpha) on skin and in saliva between the treatment groups. In addition, secondary outcome measures included differences in microbial functions in saliva and salivary cytokine levels (IL-6, IL-10) between the treatment groups. Another secondary objective was to assess whether there are associations between environmental factors, yard management practices, activities at the yard, salivary cytokine levels and bacterial measurements.

Sample size calculations for prior effect estimates are in appendix p 3.

### 2.3. Experimental design Rewilding group

Private yards of the rewilding group were modified with berry bushes, fruit trees, perennial yard plants, meadow transplants (1 m^3^), cultivation boxes (n = 3), organic mulch materials, decaying deadwood, leaf compost and organic soil with high microbial diversity in July 2022. The planting soil had high microbial richness.^10,35,36^ It was manufactured from composted materials (wood fiber, chicken manure, bark grit, *Sphagnum* moss, biochar of vegetable origin, light growing peat; manufacturer Biolan Oy, Eura, Finland; trade names Musta Multa, Puutarhan Sammalmulta, Turpeeton Puutarhamulta). Berry bushes, fruit trees and yard plants were chosen based on high microbial diversity and richness in leaf buds (Appendix p 5). Rewilding group received herbs for cultivation boxes (Appendix p 4). One planting box per yard was left empty for annual edible plants.

Decaying deadwood was transferred to yards as 1 m silver birch (*Betula pendula*) logs and wood chips (birch, aspen and oak). The logs had been inoculated with two fungi, lion’s mane (*Hericium erinaceus*) and shiitake (*Lentinuula edodes*) two years earlier by KÄÄPÄ Biotech, Lohja, Finland.

Plants, cultivation boxes and decaying deadwood were placed near walkways, doors and terraces. The rewilding group received a leaf compost and were instructed to recycle organic matter and nutrients on site, e. g., to reduce raking, chop autumn leaves onto lawn and compost plant material.

To improve the completeness of reporting, and ultimately the replicability of intervention, we used the Template for Intervention Description and Replication (TIDieR) checklist and guide that is an extension of the CONSORT 2010 statement.^37^

Intervention lasted 90 days from July 2022 to November 2022.

### Control group

The control group’s yards were not modified. Control group received inorganic fertilizers, moss remover and ant control insecticide (Myrr®; active ingredient imidacloprid 0.03 % w/w).

### 2.4. Skin swab collection and microbial analyses

Skin swabs were collected at baseline in July 2022 and three months later in early November 2022. Skin swab samples were collected with a sterile cotton-wool stick wetted in 0.1% Tween® 20 in 0.15 M NaCl from two parts of the dominant hand: 1) back of the hand (4 × 4 cm area), 2) forearm (5 × 5 cm area) (10 seconds wiping). Skin swabs were stored at -20 ◦C and within a week transferred to -80 ◦C.

DNA was extracted with DNeasy® PowerSoil® Pro Kit (Qiagen) according to manufacturer’s standard protocol. PCR were performed for the V4 region within the 16S rRNA gene (three technical replicates from each sample) using 505F and 806R primers.^38^ Negative controls (sterile water) and the ZymoBIOMICS^TM^ Microbial Community Standard (Zymo Research Corp., Irvine, CA, United States) were included in DNA extraction and PCR to ensure the quality of the analysis. Paired-end sequencing of the amplicons (2 X 300 bp) was performed on an Illumina Miseq instrument using a v3 reagent kit.

Raw paired-end sequence files were processed into amplicon sequence variants (ASVs) using DADA2^39^ with the non-redundant Silva database version 138.^40^ To conceptualize the different sequence read depths, the data were transformed to proportions by dividing the reads for each amplicon sequence variant (ASV) in a sample by the total number of reads in that sample.^41^

### 2.5. Saliva sample collection, metagenome shotgun sequencing and cytokine measurements

Saliva samples for microbial analyses were taken in the morning immediately after waking up. The participants held three sterile cotton swabs in mouth for 40s. Thereafter, saliva samples for cytokine analysis were collected into empty collection tubes with no additives by passive drooling before eating or brushing teeth. The samples were subsequently frozen at -20 ◦C and within a week transferred to -80 ◦C.

Whole metagenome shotgun sequencing was performed at FIMM – Institute for Molecular Medicine Finland as part of the Helsinki Institute of Life Science HiLIFE at the University of Helsinki. Libraries generated from total genomic DNA extracted from saliva was sequenced on the Noveseq S1/300c (Illumina) platform using the 2x150 bp paired-end read protocol. The quality of the sequencing reads was assessed with FastQC and low-quality, ambiguous sequences, adapters and contaminants were removed. Taxonomic profiling of the final read set was performed using Kraken2,^42^ while functional profiling was carried out using HUMAnN3.^43^

For cytokine measurements, samples were thawed and IL-6 and IL-10 concentrations determined with a custom U-plex kit and MESO Quickplex SQ 120 by Meso Scale Discovery (Rockville, MD, US), according to manufacturer’s instructions. Samples were used undiluted.

### 2.6. Land cover category classification

The land cover categories surrounding the dwellings of the participants were analyzed with the preclassified Coordination of Information on the Environment (CORINE) Land Cover 20-m raster.^44^

The land cover analysis was performed using the data from the year 2018. The calculations used in this study included three buffer-zones, 500, 1000, 1500-meter radii around dwelling coordinates.

### 2.7. Vegetation, deadwood and polypore fungi inventories

A vegetation inventory was done in each private yard and background information of the yard was recorded at baseline (June-July) (Appendix p 3). Deadwood and fruiting-bodies of wood-inhabiting polypore fungi^28^ inventories were done within 200 m radii around the yards. Deadwood inventories encompassed all green areas and trees within public areas. We inventoried all dead trees with ≥ 15 cm diameter at breast height (1.3 m) and other pieces of coarse woody debris with ≥ 15 cm basal diameter and ≥ 1.3 m length. For each dead tree and piece of deadwood, we recorded tree species and decay class^45^ and occurrences of polypore fruiting-bodies. In addition, polypore fruiting-bodies were recorded from living trees with internal decay or attached dead branches. Decaying deadwood in our study represents decay class ≥ 2 in Renvall (1995) classification: Wood fairly hard and knife penetrates ca. 1-2 cm into the wood.^45^

### 2.8. Statistical analyses

All the statistical tests were done with R v4.3.1^46^ and with *vegan*^47^ and *lme4*^48^ packages. Linear Mixed-effect Models (LMM) (function *lmer* in *lme4* package) was constructed to analyze temporal shifts in microbial measurements and correlation between microbial variables and living environment (Appendix p 4). Linear Discriminant Analysis Effect Size (LEfSe) was used to test correlations between the amount of deadwood and fungi polypore, and skin microbial taxonomies. The changes and difference between treatments in questionnaire data were determined using generalized linear models (GLM) with binomial distributions [function *glmer* in *lme4* package]. Correlations between yard characteristics and salivary cytokine levels were determined using GLM with quasipoisson distributions.

Skin and salivary microbial beta diversity was analyzed with Permutational Multivariate Analysis of Covariance (PERMANOVA, function *adonis2* in *vegan* package) with Bray-Curtis metric (Appendix p 4).^49^ To test the correlation between beta diversity of skin and salivary microbiota and living environment, principal coordinates analysis (PCoA) with Bray-Curtis distance was used to score the microbial measurements onto an ordination and correlation with environmental factors was assessed using function *envfit* in *vegan* package.

To verify the P-value approximations, permutation tests (*lmperm* in *permuco* package) were run (5,000 permutations).^50^ All the statistical tests were considered significant when permuted or Benjamini-Hochberg adjusted p-value was < 0.05 level.

## 3. Results

### 3.1. Characteristics of yards and study subjects were similar in both treatment groups

Thirty-six healthy volunteers aged between 33–70 participated in the study (Table 1). Major land cover category around all yards was artificial surfaces, particularly discontinuous urban fabric (Table 1). This was true for all radiuses (100-2500 m). Yard characteristics were similar between treatment groups, except that the control group had approximately 16% more of manicured yard area (P = 0.05; Appendix p 5). According to the questionnaires, gardening was practiced as often, i.e., at least weekly, in both groups during the study period. The study groups did not differ in the amount of decaying deadwood and polypore species richness around homes (P > 0.1). At the baseline, yard management practices, activities in the yard, nature visits, and animal and soil contacts were similar between treatment groups (P > 0.05).

During the study period, rewilding group had planted more often as compared to the control group (GLM: P = 0.002, R2 = 0.22; Appendix p 6). None of the participants had used plant pesticides. Insecticides were used rarely or never. Rewilding group did not use inorganic fertilizers during the study period, whereas among control group these were used 1-3 times in a month or more rarely. Time spent at the home yards decreased in fall in both the intervention and control groups (Appendix p 6-7).

### 3.2. Neighborhood deadwood and land cover were associated with commensal microbiota

LEfSe analysis (Figure 1) and LMM models both identified Proteobacteria, particularly gammaproteobacterial order Burkholderiales and family Neisseriaceae, on the forearm to be associated with high polypore species richness and the amount of decaying deadwood (P < 0.001; Appendix p 8). In addition, skin bacterial beta diversity was associated with deadwood both at the baseline (forearm P = 0.024, R2 = 0.22; Figure 2B) and in the fall (back of the hand P = 0.035, R2 = 0.30; Figure 2A) (Appendix p 9).

**Figure 1:**
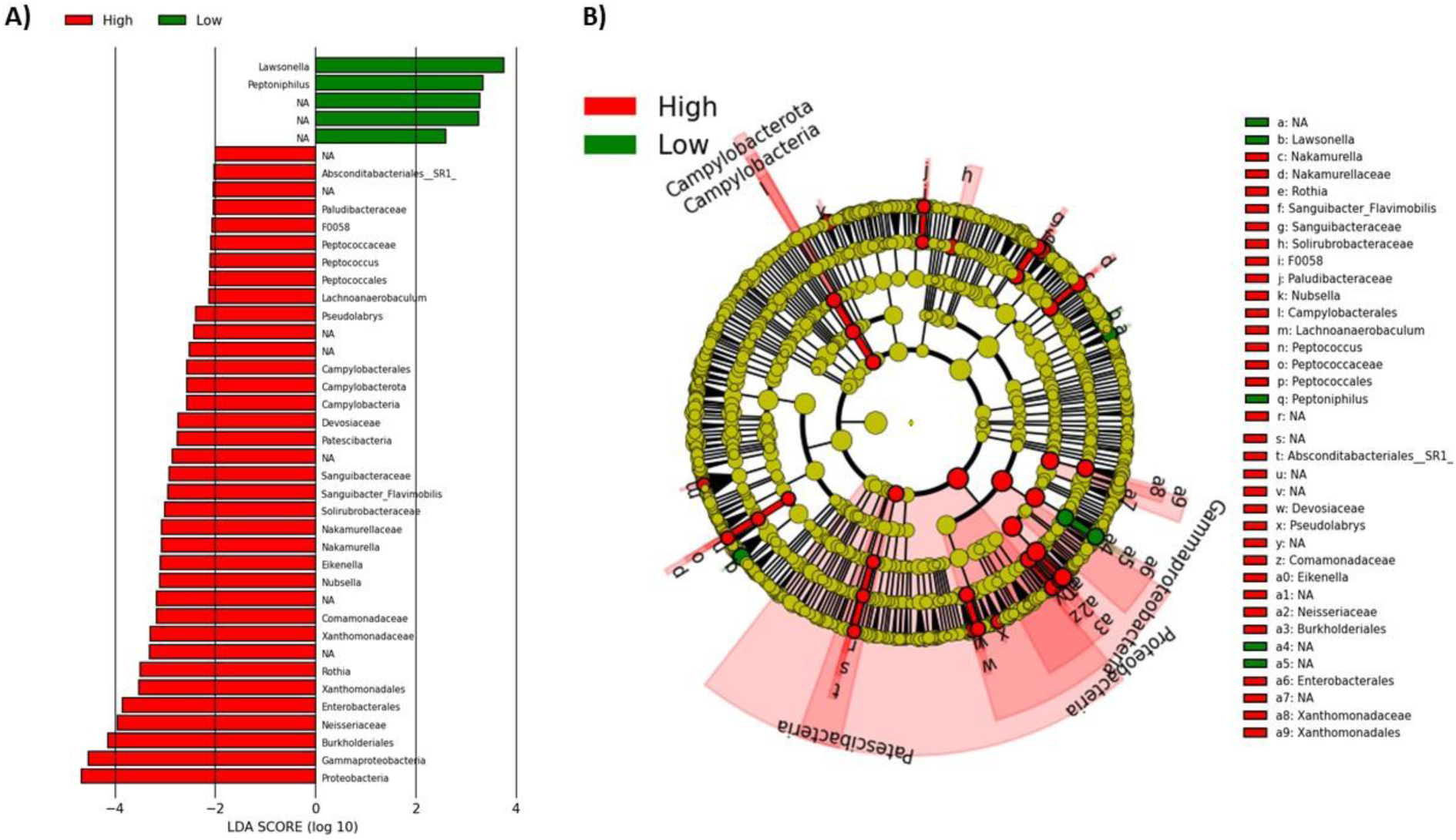
Linear discriminant analysis effect size (LEfSe) analysis of skin bacterial abundance on the arm of yard inhabitants. The study subjects were split to two groups based on the amount of neighborhood deadwood: High (red) > 10 and low (green) < 10 deadwood logs. A) Linear discriminant analysis (LDA) score identified the size of differentiation with a threshold value of 2.0. B) The cladogram of skin microbial communities on the arm. Unclassified bacteria = NA.

**Figure 2.**
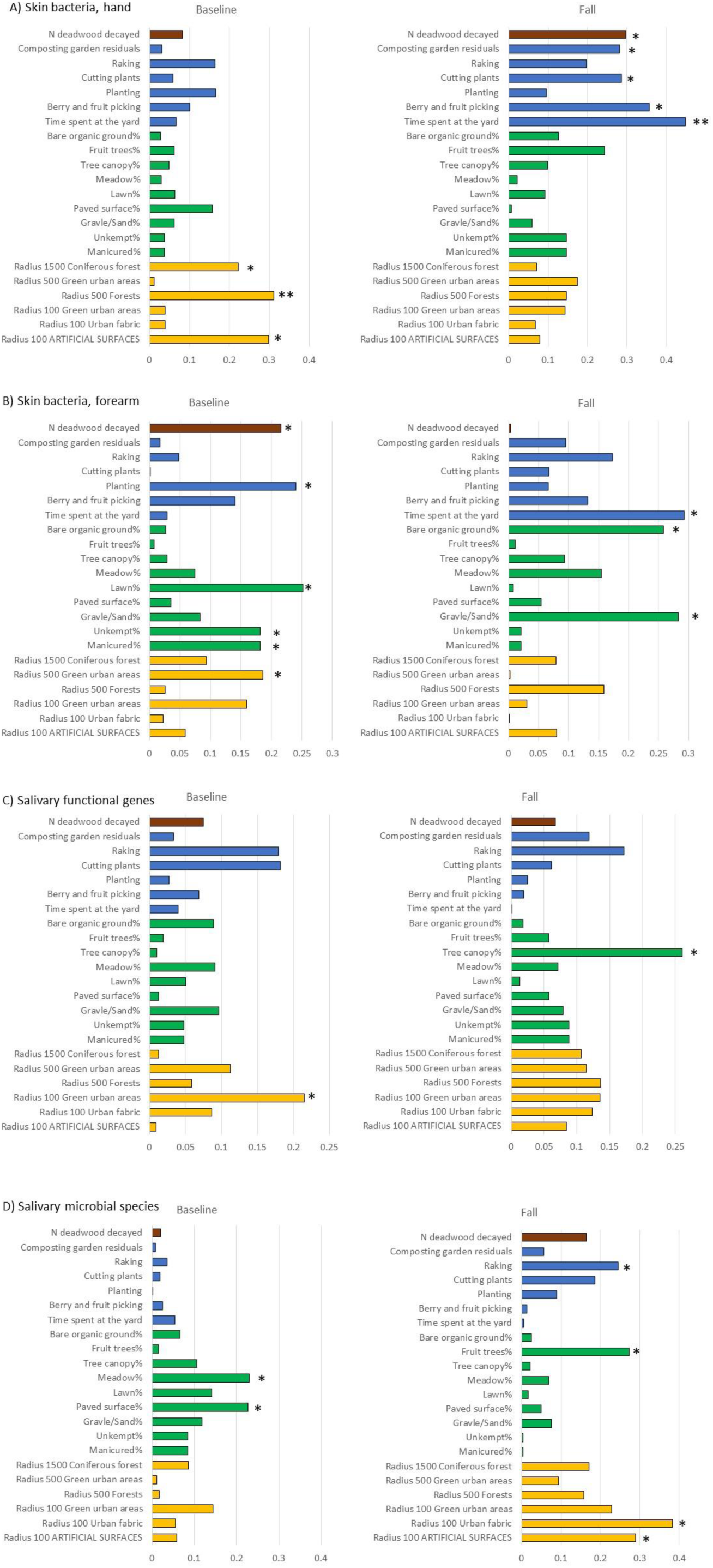
Significance and explained variance of 22 environmental variables modelled by EnvFit. Horizontal bars show the amount of variance (R2) explained by each environmental variable. A) skin bacteria on the back of the hand and B) on forearm are based on 16S rRNA sequencing data at ASV level, and C) salivary functional gene pathways and D) salivary microbial species are based on metagenomic shotgun sequencing data at baseline and in fall (day 90). Environmental variables are colored based on metadata group: brown = deadwood, blue = garden management practices, green = yard characteristics (percentage of yard area), yellow = land cover categories around home. * P < 0.05, ** P < 0.01.

In saliva, Simpson diversity index of functional gene pathways correlated positively with the amount of neighborhood deadwood, but only at baseline (LMM: P = 0.04, R2 = 0.12). City was a significant confounding factor in LMM models because Helsinki had more deadwood and higher polypore species richness compared to Kaarina (P < 0.04) and Lahti (P < 0.0001).

At the baseline, beta diversity of functional gene pathways in saliva was associated with green urban areas at 100-m radius (P = 0.019, R2 = 0.21; Figure 2C). Beta diversity of skin bacterial taxa on the back of the hand was associated with forests (P = 0.007, R2 = 0.31), while on the forearm it was associated with green urban areas at 500-m radius (P = 0.04, R2 = 0.19) (Figure 2A and B). In fall these associations were not observed (P ≥ 0.069).

### 3.3. Yard management practices and yard characteristics influenced the abundance profile of salivary and skin microbiota

At the baseline, the coverage of paved surfaces (P = 0.027, R2 = 0.12) and meadows (P = 0.03, R2 = 0.23) at the yard were associated with the abundance profile of salivary microbial species (Figure 2D). In fall, the associations were not observed anymore, but urban fabric around yards with 100-radius (P = 0.012, R2 = 0.38; Figure 2D) and raking frequency (P = 0.047, R2 = 0.24; Figure 2D) influenced the abundance profile of salivary microbial species to the same direction (Appendix p 16). Control study subjects raked more often their yards in fall compared to the baseline (P = 0.02, R2 = 0.33; Appndix p 7).

The abundance profile of forearm skin bacteria depended on manicured yard area% (P = 0.042, R2 = 0.18; Figure 2B) and lawn% (P = 0.022, R2 = 0.25; Figure 2B) to the opposite direction as unkept yard area% (P = 0.042, R2 = 0.18; Appendix p 16). In fall, the frequencies of berry and fruit picking (P = 0.014), composting garden residues (P = 0.044) and cutting plants (P = 0.039) influenced skin bacterial community (back of hand) (Figure 2A; Appendix p 16). Time spent at the yard influenced skin bacteria composition in fall (Figures 2A and B).

Confounding factors including age, gender, BMI, and diet were not associated with salivary microbial profiles (Appendix p 10). Age (forearm) and junk food consumption (back of hand) were associated with skin bacterial profile (Appendix p 10).

### 3.4. Rewilding preserved skin bacterial diversity and richness to fall

Skin bacterial richness or diversity did not decrease in the rewilding group during the study period (P > 0.5), while the alphaproteobacterial Simpson diversity decreased among control group on the forearm (P = 0.008, R2 = 0.19) (Figure 3; Appendix p 12). Beta diversity of skin bacteria on the back of the hand shifted in the control group during the study period, while no changes were observed in the rewilding group (Figure 4). Skin alphaproteobacterial diversity decreased from summer to fall also when both groups were analyzed together (Hand: P = 0.003, forearm: P = 0.004; Appendix p 12-13).

**Figure 3.**
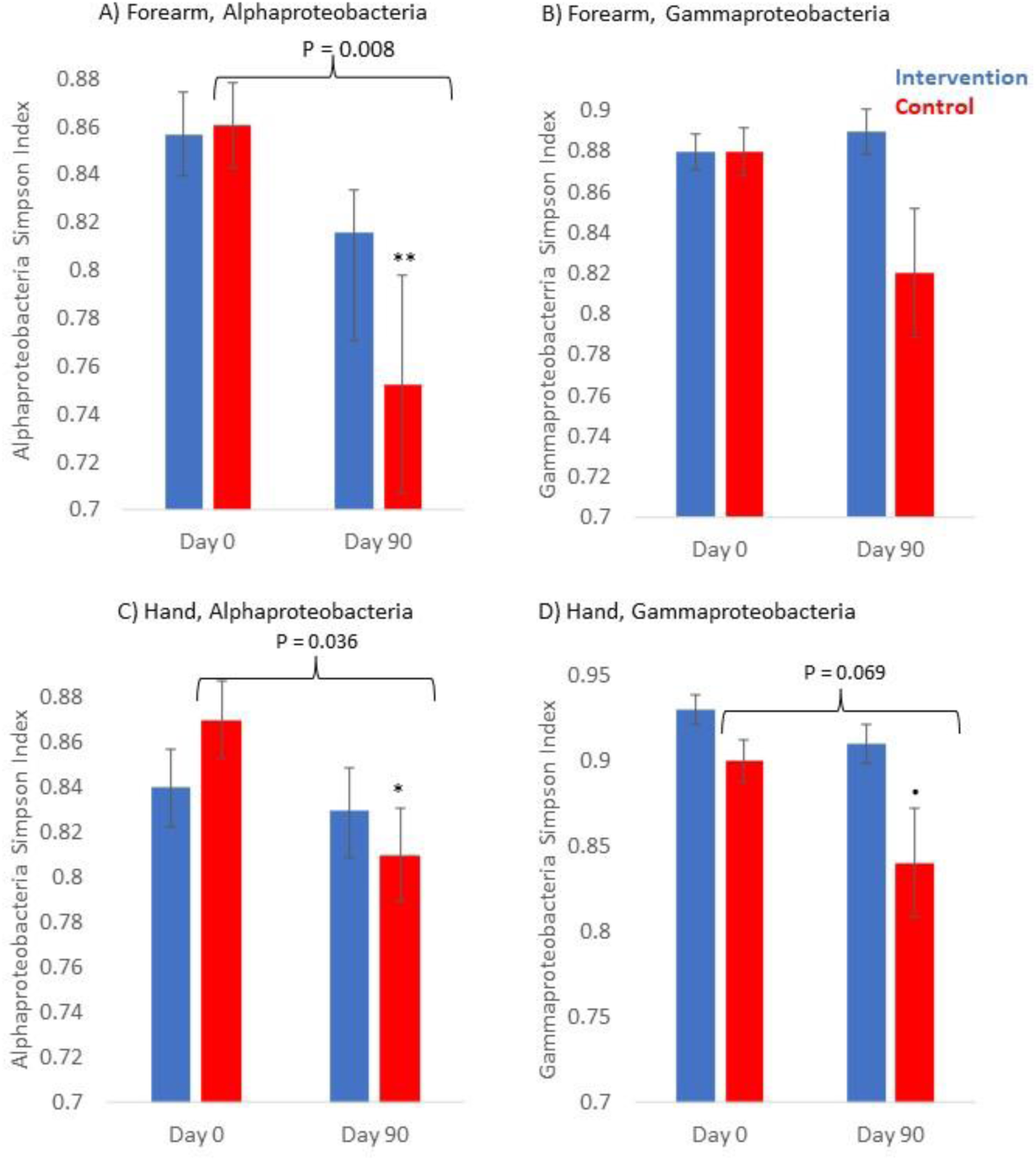
Skin Simpson diversity index of A) Alphaproteobacteria and B) Gammaproteobacteria on forearm, and C) Alphaproteobacteria and D) Gammaproteobacteria on back of the hand between intervention and control groups at baseline (Day 0) and in fall. (Day 90). Decrease in control: P = 0.07, *P = 0.04, **P = 0.008

**Figure 4.**
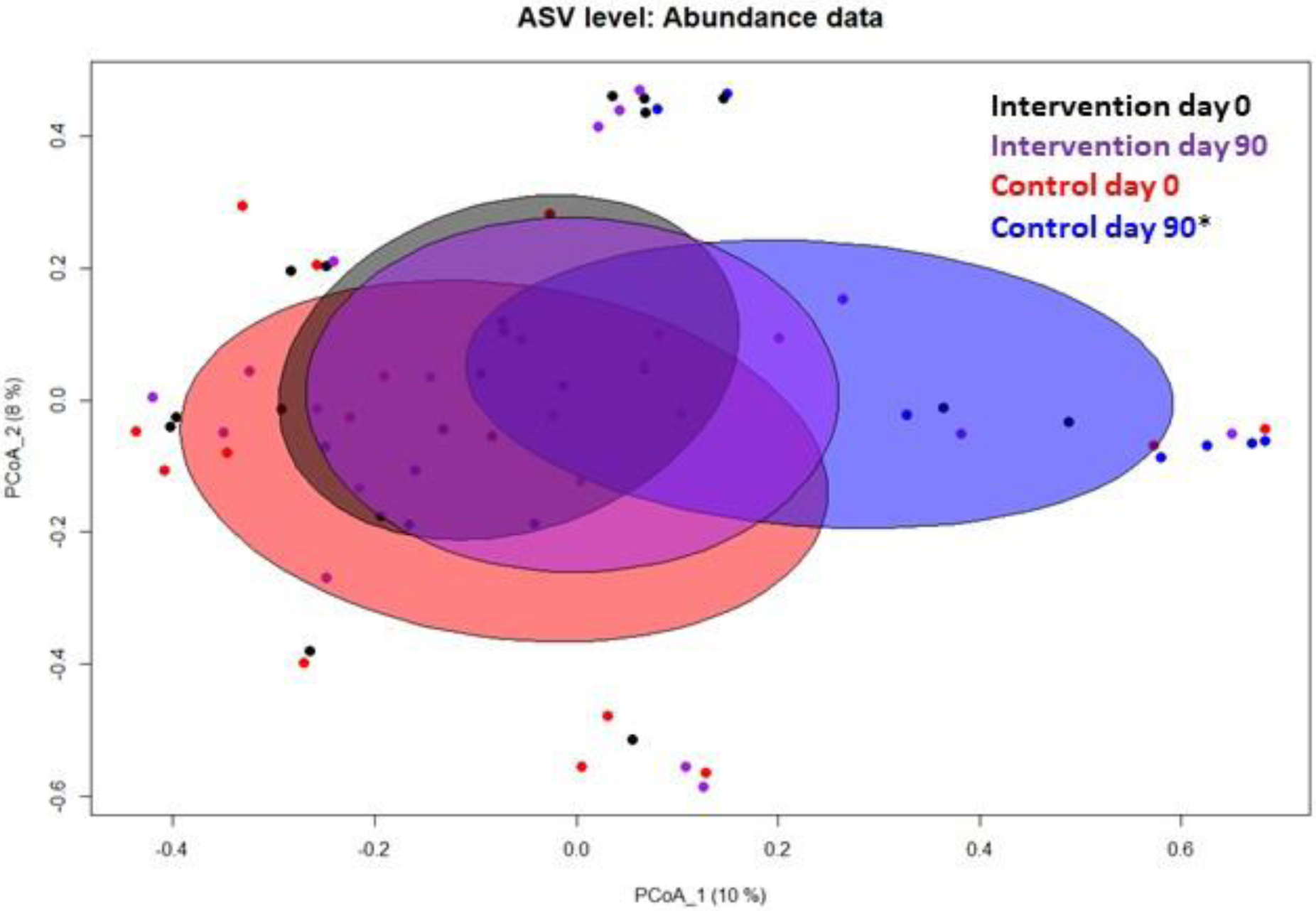
Beta diversity of skin bacteria on the back of the hand. *Composition of skin bacteria shifted among control subjects P = 0.046 (Bray-Curtis distance in PERMANOVA).

At the baseline, yard plant richness was directly associated with skin alphaproteobacterial diversity on the back of the hand (P = 0.007; Table 2A) and on the forearm (P = 0.008; Appendix p 17). In the fall, plant richness was directly associated with total bacterial richness and diversity on the back of the hand (P ≤ 0.01; Table 2A) and the richness on the forearm (P = 0.05) (Appendix p 17). When treatment groups were analyzed separately, the direct association between plant richness and skin total bacterial richness on the back of the hand (P = 0.006) and diversity (P = 0.004) were observed only within the rewilding group in the fall (Table 2B; Appendix p 17). Treatments did not differ in yard plant richness at the baseline of in fall (Appendix p 5).

**Table 2.**
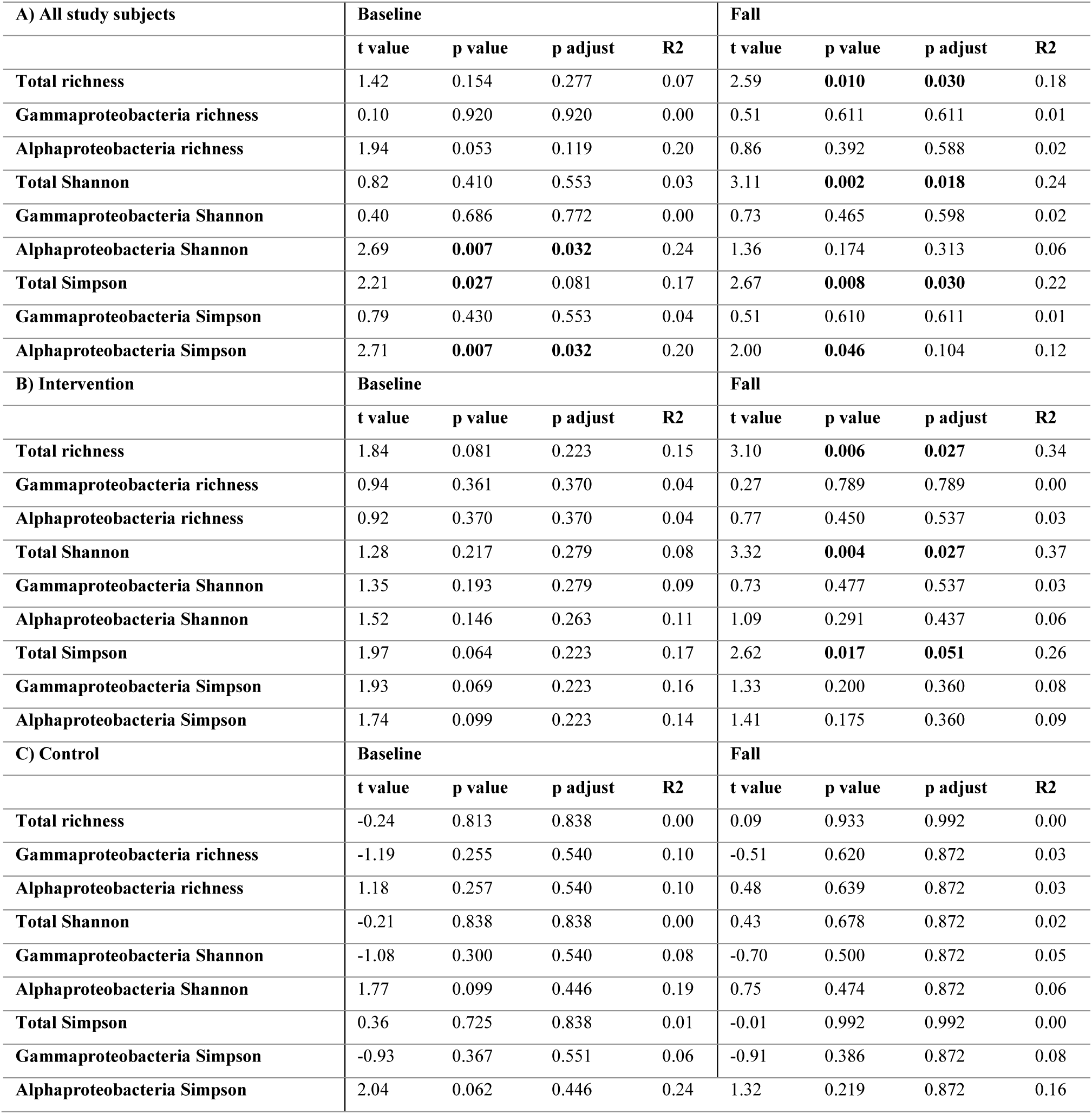
Linear mixed model (LMM) results for correlation of yard plant richness with skin bacterial richness and diversity indices on the back of the hand. Analyses were done with **A)** all study subjects, and **B)** intervention study subjects, and **C)** control study subjects at baseline and three months later in fall. LMM statistics are reported as t value, probability p value, Benajmini-Hochberg adjusted p value and R squared (R2).

| A) All study subjects | Baseline |  |  |  | Fall |  |  |  |
| --- | --- | --- | --- | --- | --- | --- | --- | --- |
|  | t value | p value | p adjust | R2 | t value | p value | p adjust | R2 |
| Total richness | 1.42 | 0.154 | 0.277 | 0.07 | 2.59 | **0.010** | **0.030** | 0.18 |
| Gammaproteobacteria richness | 0.10 | 0.920 | 0.920 | 0.00 | 0.51 | 0.611 | 0.611 | 0.01 |
| Alphaproteobacteria richness | 1.94 | 0.053 | 0.119 | 0.20 | 0.86 | 0.392 | 0.588 | 0.02 |
| Total Shannon | 0.82 | 0.410 | 0.553 | 0.03 | 3.11 | **0.002** | **0.018** | 0.24 |
| Gammaproteobacteria Shannon | 0.40 | 0.686 | 0.772 | 0.00 | 0.73 | 0.465 | 0.598 | 0.02 |
| Alphaproteobacteria Shannon | 2.69 | **0.007** | **0.032** | 0.24 | 1.36 | 0.174 | 0.313 | 0.06 |
| Total Simpson | 2.21 | **0.027** | 0.081 | 0.17 | 2.67 | **0.008** | **0.030** | 0.22 |
| Gammaproteobacteria Simpson | 0.79 | 0.430 | 0.553 | 0.04 | 0.51 | 0.610 | 0.611 | 0.01 |
| Alphaproteobacteria Simpson | 2.71 | **0.007** | **0.032** | 0.20 | 2.00 | **0.046** | 0.104 | 0.12 |
| B) Intervention | Baseline |  |  |  | Fall |  |  |  |
|  | t value | p value | p adjust | R2 | t value | p value | p adjust | R2 |
| Total richness | 1.84 | 0.081 | 0.223 | 0.15 | 3.10 | **0.006** | **0.027** | 0.34 |
| Gammaproteobacteria richness | 0.94 | 0.361 | 0.370 | 0.04 | 0.27 | 0.789 | 0.789 | 0.00 |
| Alphaproteobacteria richness | 0.92 | 0.370 | 0.370 | 0.04 | 0.77 | 0.450 | 0.537 | 0.03 |
| Total Shannon | 1.28 | 0.217 | 0.279 | 0.08 | 3.32 | **0.004** | **0.027** | 0.37 |
| Gammaproteobacteria Shannon | 1.35 | 0.193 | 0.279 | 0.09 | 0.73 | 0.477 | 0.537 | 0.03 |
| Alphaproteobacteria Shannon | 1.52 | 0.146 | 0.263 | 0.11 | 1.09 | 0.291 | 0.437 | 0.06 |
| Total Simpson | 1.97 | 0.064 | 0.223 | 0.17 | 2.62 | **0.017** | **0.051** | 0.26 |
| Gammaproteobacteria Simpson | 1.93 | 0.069 | 0.223 | 0.16 | 1.33 | 0.200 | 0.360 | 0.08 |
| Alphaproteobacteria Simpson | 1.74 | 0.099 | 0.223 | 0.14 | 1.41 | 0.175 | 0.360 | 0.09 |
| C) Control | Baseline |  |  |  | Fall |  |  |  |
|  | t value | p value | p adjust | R2 | t value | p value | p adjust | R2 |
| Total richness | -0.24 | 0.813 | 0.838 | 0.00 | 0.09 | 0.933 | 0.992 | 0.00 |
| Gammaproteobacteria richness | -1.19 | 0.255 | 0.540 | 0.10 | -0.51 | 0.620 | 0.872 | 0.03 |
| Alphaproteobacteria richness | 1.18 | 0.257 | 0.540 | 0.10 | 0.48 | 0.639 | 0.872 | 0.03 |
| Total Shannon | -0.21 | 0.838 | 0.838 | 0.00 | 0.43 | 0.678 | 0.872 | 0.02 |
| Gammaproteobacteria Shannon | -1.08 | 0.300 | 0.540 | 0.08 | -0.70 | 0.500 | 0.872 | 0.05 |
| Alphaproteobacteria Shannon | 1.77 | 0.099 | 0.446 | 0.19 | 0.75 | 0.474 | 0.872 | 0.06 |
| Total Simpson | 0.36 | 0.725 | 0.838 | 0.01 | -0.01 | 0.992 | 0.992 | 0.00 |
| Gammaproteobacteria Simpson | -0.93 | 0.367 | 0.551 | 0.06 | -0.91 | 0.386 | 0.872 | 0.08 |
| Alphaproteobacteria Simpson | 2.04 | 0.062 | 0.446 | 0.24 | 1.32 | 0.219 | 0.872 | 0.16 |

At the baseline, yard sport activities were directly associated with skin alphaproteobacterial richness (GLM: P = 0.005, R2 = 0.25) and Shannon diversity (GLM: P = 0.007, R2 = 0.24). When treatment groups were separate, berry and fruit picking was directly associated with skin bacterial richness (P = 0.01, R2 = 0.42) and Shannon diversity (P = 0.03, R2 = 0.23) in the rewilding group. In the control group, the frequency of nature visits increased skin gammaproteobacterial diversity in the fall (P = 0.002, R2 = 0.54).

### 3.5. Rewilding shifted salivary microbiota and functional gene pathways

Among rewilding group, salivary microbiomes were enriched with the family Mycobacteriaceae (P = 0.04; R2 = 0.09) and genus *Leuconostoc* (Lactobacillaceae) (P = 0.02; R2 = 0.12; Appendix p 14). Among control subjects, the relative abundance of these taxonomies showed a decreasing trend (Appendix p 14). The change difference between treatments was significant (LMM: P = 0.007, R2 = 0.20 and P = 0.018, R2 = 0.15, respectively; Appendix p 14). Functional gene pathway related to L-histidine degradation I was decreased during the study period in the rewilding group, while it increased in the control group (Change difference between treatments in LMM: P = 0.006, R2 = 0.13; Appendix p 14).

### 3.6. IL-6 was associated negatively with salivary microbial and functional richness

In the fall, total microbial, alpha- and gammaproteobacterial richness and functional richness and diversity of gene pathways in saliva were negatively associated with salivary IL-6 (Table 3; Appendix p 18). Picking of berries and fruits were directly associated with salivary IL-10 (P = 0.002) and IL-6 (P = 0.006) (Appendix p 15) in the rewilding group but not in the control group or at the baseline (P > 0.1; Appendix p 15). Fruit trees and tree canopy percentage were directly associated with salivary IL-10 (fruit trees% P = 0.015) and IL-6 (fruit trees% P < 0.001 and tree canopy% P = 0.002) at the baseline (Appendix p 15). Potential confounding factors (age, gender, BMI and diet) were not associated with immune markers (Appendix p 10-11).

**Table 3.** Linear mixed model (LMM) results in fall between salivary IL-6 and microbial richness and diversity indexes. LMM statistics are reported as t value, probability p value, permuted p value (5,000 permutaitons), Benjamini-Hochberg adjusted p value and fixed R squared (R2) for the model.

|  | **t value** | **p value** | **permuted p value** | **p adjust** | **R2** |
| --- | --- | --- | --- | --- | --- |
| Total richness | -3.076 | **0.005** | **0.003** | **0.017** | 0.273 |
| Phyla Proteobacteria richness | -2.750 | **0.010** | **0.004** | **0.017** | 0.251 |
| Class Gammaproteobacteria richness | -2.501 | **0.019** | **0.007** | **0.017** | 0.229 |
| Class Alphaproteobacteria richness | -3.225 | **0.003** | **0.003** | **0.017** | 0.278 |
| functional richness | -2.910 | **0.007** | **0.005** | **0.017** | 0.255 |
| Total Shannon | -2.223 | **0.035** | **0.032** | 0.054 | 0.158 |
| Phyla Proteobacteria Shannon | -0.679 | 0.503 | 0.515 | 0.594 | 0.016 |
| Class Gammaproteobacteria Shannon | -0.923 | 0.364 | 0.375 | 0.468 | 0.030 |
| Class Alphaproteobacteria Shannon | -2.797 | **0.009** | **0.009** | **0.020** | 0.246 |
| Functional Shannon | -2.894 | **0.007** | **0.006** | **0.017** | 0.245 |
| Total Simpson | -0.993 | 0.330 | 0.318 | 0.433 | 0.036 |
| Phyla Proteobacteria Simpson | 0.147 | 0.884 | 0.891 | 0.891 | 0.001 |
| Class Gammaproteobacteria Simpson | -0.559 | 0.581 | 0.612 | 0.656 | 0.011 |
| Class Alphaproteobacteria Simpson | -1.232 | 0.228 | 0.147 | 0.220 | 0.057 |
| Functional Simpson | -2.474 | **0.020** | **0.013** | **0.025** | 0.210 |

## 4. Discussion

### 4.1. Yard rewilding promotes health-promoting ecosystem services

Our study is the first to show how rewilding of urban private yards with vegetation and decaying deadwood prevents the microbial diversity loss on human skin and plausibly modifies salivary microbial community, functional gene pathways, and immune response. Our results indicate that deadwood and plant litter are at least as crucial as plant richness for the ecosystem services provided by green space.

Several factors underscore the significance of rewilding as a health-enhancing ecosystem service in our study. First, the difference between the two treatments in diversity and richness of skin proteobacterial classes were observed, even though gardening was practiced with similar frequency, and time spent at the yards decreased in autumn in both groups. Second, since we specifically selected plant cultivars hosting rich bacterial, and particularly gammaproteobacterial community, these cultivars might have contributed to the observed difference. The same applies to gardening advice provided by our team; the rewilding group left plant litter to decompose naturally, which increases soil microbial diversity and resilience, and enhances nutrient cycle *on site.*^51,52^ The use of logs and chips, and biodiverse planting soil were designed to increase the overall and Gammaproteobacterial exposure at the yards, and thus could contribute to the difference between the two study groups; Gammaproteobacteria have been linked to improved immune regulation and a reduced atopy risk.^2,19,53^ In earlier intervention trials,^19,53^ these bacterial classes have been associated with the higher frequency of T regulatory cells, which play a critical role in preventing autoimmunity.^54^ Therefore, our finding indicates improved homeostasis and immunological self-tolerance.

Differences in the consequences of living habits also support the importance of rewilding. Only within the rewilding group picking of edible plants at the yard was associated with skin microbial diversity. In line with this, the frequency of nature visits increased skin gammaproteobacterial diversity within the control group only. Since we added several elements that plausibly enhanced proteobacterial community on the skin, it is hard to distinguish the core reason for the differences in skin microbiota between intervention and control groups. Nevertheless, our findings underline the importance of the quality of urban green space. Additional research is evidently needed to reveal the relative importance of each element. Today, the viable solution provided by the current study is to optimize them all.

### 4.2. Neighborhood deadwood promotes health-associated skin bacteriome

The hypothesis that decaying wood is an important factor modulating skin microbiota of urbanities is supported by our study. First, proteobacterial taxa on the skin were directly associated with the decaying deadwood in the living environment. Since decaying deadwood exists in boreal nature together with other decomposing plant matter, and a vivid moss layer, the finding highlights the importance of plant litter in general. Interestingly, the same bacterial taxa that we found abundantly on skin, i.e., Alpha- and Gammaproteobacteria and Burkholderiales, are rich in decaying wood.^55–60^

Among the most interesting findings of our study were the associations between gardening practice and land cover. While the coverage of unkept garden area and deadwood shaped skin microbial community to an opposite direction than a high coverage of manicured lawn, frequent raking and urban fabric, including asphalt, shaped salivary microbial community similarly (Appendix p 16). This indicates that the removal of autumn leaves and other plant litter modifies the commensal microbiome to the same direction as built-up urban area. In several earlier studies,^2,27^ buildings and paved surface were associated with an increased risk of immune-mediated diseases. The current study thus provides an explanation how paving and removal of plant litter are connected to human and planetary health.

Even though previous studies pointed out the importance of vegetation for health-associated microbiota transferred indoors,^61–63^ our study is the first to separate the roles of plant richness versus deadwood abundance. Deadwood and plant residues seem to play an equal or even more important role in the health-associated ecosystem services than live vegetation. Until now, vegetation indices were a major if not the sole indicator of biodiversity in studies that tested how biodiversity is related to immune-mediated diseases.^21^ Our study opens a new avenue for research; studies focusing on the health-associated ecosystem services should investigate neighborhood deadwood and plant residues in parallel live vegetation.

### 4.3. Changes in salivary microbiota, functional gene pathways and cytokines are in accordance with the biodiversity hypothesis

The shifts observed in the salivary microbiota, functional gene pathways and cytokines are encouraging in the context of the biodiversity hypothesis. Among intervention participants, Mycobacteriaceae abundance in saliva increased; soil-derived *Mycobacterium* reduced inflammation and enhanced stress-resilience in murine studies.^64–66^ Because Mycobacteriaceae prevails in decaying deadwood,^67^ the increase in the rewilding group may be due to the deadwood logs and chips used in the rewilding coupled with microbially diverse soil for the planting boxes. In parallel with Mycobacteriaceae, the observed increase in *Leuconostoc* sp. in saliva among the intervention participants may have metabolic benefits, because this probiotic genus is well known for fermentative potential.^68,69^

The analysis of salivary gene pathways indicates immunomodulatory benefits of the intervention treatment. Since the degradation of L-histidine is interconnected with various metabolic pathways that are involved in the biosynthesis of essential amino acids,^70^ the observed shifts in L-histidine degradation pathway in saliva may have implications for cellular function, neurotransmission, immune regulation, and overall physiological balance and well-being. Because L-histidine degradation pathway I degrades L-histidine to L-glutamate, and because L-glutamate is associated with chronic inflammation^71,72^, the decrease in this pathway should contribute to reduction of pro-inflammatory signals. A recent in *vitro study* suggested that L-histidine attenuates non-esterified fatty acids -induced inflammation by reducing Grb2-associated binder 2 expression, and eventually induces IL-6 expression.^73^ In our study, the abundance of decaying deadwood diversified salivary gene pathways, which in turn decreased IL-6 levels in saliva. Higher pro-inflammatory salivary cytokine levels have been associated with immune-mediated diseases, periodontal and tumor diseases,^74,75^ stress^76,77^ and neuropsychiatric disorders.^78^ Further support for the immunomodulatory role of rewilding comes from the observation that picking edible plants was associated with salivary cytokines in the rewilding group. In other words, the variables that enhanced skin microbial richness and diversity were also associated directly with salivary cytokines only among rewilding group. As a whole, microbiologically oriented rewilding may be beneficial to mental and physical health.

## 5. Conclusions

This study showed that urban rewilding induces immunomodulatory shifts in salivary microbiota, functional gene pathways and cytokine levels, and that decaying deadwood induces beneficial shifts on the skin microbiota at least as efficiently as plant richness. The importance of the biologically diverse living environment in immunomodulation underscores the need to apply a microbially oriented rewilding approach in urban green spaces. Future studies should use manually inventoried and satellite-data-based deadwood/plant litter indices in parallel with vegetation indices when estimating the associations between biodiversity and immune-mediated diseases. Our findings provide an incentive for future urban rewilding approaches with microbially enriched organic soil, plants, and decaying deadwood for planetary health.

## Acknowledgements

We thank all BIWE team members and molecular laboratory personnel Katri Leino and Tuija Hytönen, and specialist Leena Vuorinen at Natural Resources Institute Finland, and trainee Hibak Weiss at Tampere University. A special thank-you are owed to the study participants for their participation in this study. The authors wish to acknowledge CSC – IT Center for Science, Finland, for computational resources, and FIMM Genomics NGS Sequencing unit at University of Helsinki supported by HiLIFE and Biocenter Finland.

## Author contributions

A.S., M.I.R., R.P., L.U., A.O., L.K., J.R, O.H.L. designed study; M.I.R., L.U., A.O., A.L., E.H., A.S., J.R. and O.H.L. implemented study; M.I.R., A.O., L.K., A.L., D.C., A.K., R.P., and E.H. generated data; M.I.R. and A.S. analyzed data; M.I.R. wrote first draft of the manuscript and prepared the figures and tables; All the authors reviewed the manuscript; H.H. was the medical monitor; A.S. was the principal investigators of the project.

## Data availability

Skin bacterial sequence data were accessioned into the Sequence Read Archive (BioProject ID: PRJNA1102262). All other data needed to support the conclusions of this manuscript are included in the main text and supplementary appendix.

The sensitive data that support the findings of this study are available from Natural Resources Institute Finland, but restrictions defined in General Data Protection Regulation (EU 2016/679) and Finnish Data Protection Act 1050/2018 apply to the availability of these data, and so are not publicly available. Data are however available from the authors upon reasonable request and with permission from the ethical committee of the local hospital district (Helsinki and Uusimaa Hospital District, Finland).

